# Philosophy of Maternity Nursing: Women Centered Care

**DOI:** 10.1101/2022.10.06.22280764

**Authors:** Aria Aulia Nastiti, Moses Glorino Rumambo Pandin, Nursalam

## Abstract

**Introduction:** Maternity nursing is a discipline part of nursing that focuses on the degree of health of women in this stage of life as the main object. Various women’s health problems that occur at this time along with the rapid development of technology have become a challenge for maternity nursing as a comprehensive care service to continue to develop for the better and collaborate with other sciences. The purpose of this paper is to explain the current conditions in Indonesia and the world regarding maternity nursing that supports women’s health and quality of life.

**Method:** This article was compiled through a literature search for the authenticity of this study using articles in English, Sciencedirect, PubMed, Scopus, Research Gate, and Google Scholar from 2015 to 2022.

**Result:** The results of the literature search show that currently, maternity nursing continues to develop and is centered on women at every stage of life. Research is currently being carried out on women’s welfare, autonomy and women’s empowerment, such as the decision to use contraception and the best care during pregnancy and childbirth.

## 1. Introduction

Maternity nursing is professional nursing care given to individuals and community groups centered on women’s health in both healthy and sick conditions covering physical and psychosocial aspects at every stage of life (*life cycle approach*) from toddlers, adolescents, reproductive age to post-menopausal which consists of reproductive health (gynecology) and obstetrics (obstetrics). Similar to others, maternity nursing care provided to clients include promotive, preventive, curative, and rehabilitative. It is important to provide high-quality care to improve maternal and infant health (Koblinsky *et al*., 2016; Miller *et al*., 2016). WHO has standardized aspects of the quality of care for mothers and newborns specifically consisting of the provision of care” and “care experience” and respecting women’s autonomy during childbirth (*WHO recommendations on antenatal care for a positive pregnancy experience*, no date; Tunçalp *et al*., 2015).

Maternity nursing is an important branch of nursing science because nursing care is centered on women which in turn can have an impact on the health of the next generation. After all, healthy children will be born to healthy mothers, so maternity nurses hold a strategic key to the health and knowledge of mothers about the health of themselves and their families. For example, in the case of *stunting prevention*, maternity nursing care plays an important role, starting when adolescent girls start puberty with promotive and preventive efforts related to adequate nutrition, free from anemia, menstrual hygiene management, and being able to maintain the health of their reproductive organs to prevent early marriage. Furthermore, when women of childbearing age will marry and prepare for pregnancy, they are given nursing care related to nutrition and health screening in preparation for pregnancy. The next stage is when a woman is pregnant, nursing care is given during ante-natal care, intra-natal care, and postnatal care, including breastfeeding and nutrition for toddlers and family planning. When all of the above are given optimally, it is hoped that stunting in children will not occur.

“Women-centred care” is a term used to describe a philosophy of maternity care that promotes a holistic approach by recognizing the social, emotional, physical, spiritual and cultural needs of each woman. Expectations and contexts are determined by the woman herself (Stockman *et al*., 2014). The basic principles of woman-centred care ensure a focus on pregnancy and childbirth as the start of family life, not just as isolated clinical episodes. These phases of motherhood take full account of the meaning and value of each woman (Afulani *et al*., 2021). Women-centred care in a clinical setting is safe, supportive, and gentle. It is the philosophical foundation of maternity nursing.

Maternity nursing care is influenced by values, attitudes, and culture in society. Indonesia, which is a country with diverse cultures, considers and assesses the role of women in different regions, but all agree that pregnancy is a gift and the beginning of the process of human life, so it is important to pay attention to, as well as become a deep experience for pregnant women, their families. and society. As we know every culture there are various kinds of taboos and requirements that women must adhere to when pregnant, whether it provides health benefits or harms health. It is also a challenge how maternity nursing care is provided by taking into account the culture adopted and encouraging research related to aspects of culture and maternal health.

Postpartum period is identical with difficult times after childbirth because during this period mother faced with pain after delivery, less need sleep and stress in terms of the care of baby. Generally, postpartum maternal care involves vulva hygiene, breast care and newborn infant care. A maternity nurse can provide advice to anticipate such as to manage fatigue, improve the sleep quality and to reduce anxiety with strategy an approach that is easily accepted.

In the current condition when technology is developing very quickly, maternity nursing is also required to follow both scientifically and practically based on research and the latest situation while still paying attention to legal and ethical aspects. Shifts in roles, gender, and global and national policies related to women today also affect when women have lifestyle choices and the right to make decisions and have a high life expectancy and a good quality of life. In overcoming this, all maternity nurses must think critically, be willing to change, practice collaborative science and be quick to adapt to provide the best nursing care starting in the education (curriculum) stage to clinical and community services.

## 2. Method

The literature search for the authenticity of this study used articles in English from *Sciencedirect, Pubmed, Scopus, Research Gate, and Google Scholar* from 2015 to 2022. The literature search used the search terms: (“philosophy”) AND (“nursing “) AND (“maternity “ OR “maternal”) AND (“women centered care”)

## 3. Results

A total of 8 articles were found according to the keywords used and then analyzed. The search was performed based on the Preferred Reporting Items for Systematic Reviews and Meta-Analyses (PRISMA) guidelines (Figure 1). While the articles analyzed are shown in table 1.

**Figure 1.**
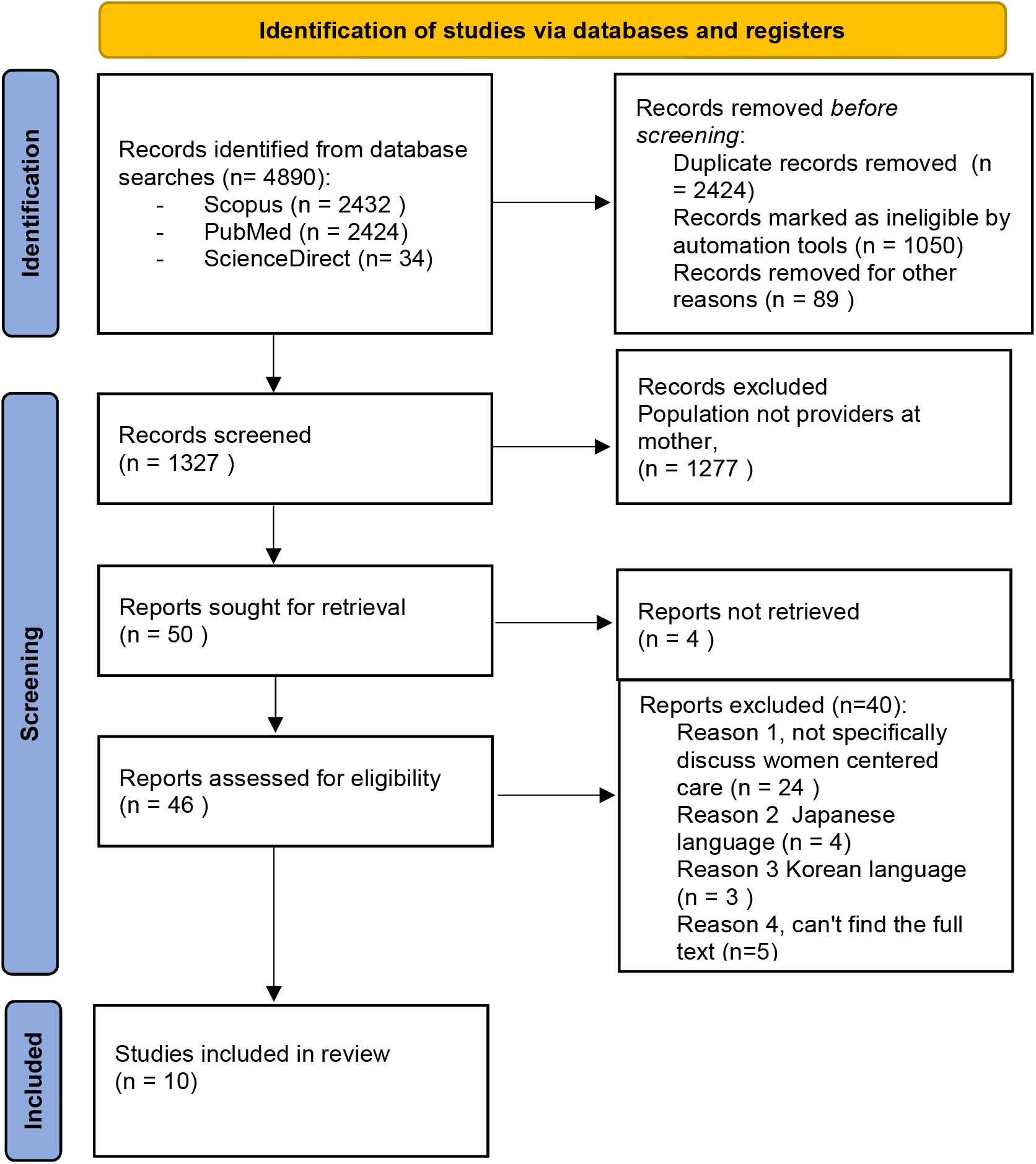
PRISMA Guideline Article Search

**Table 1.**
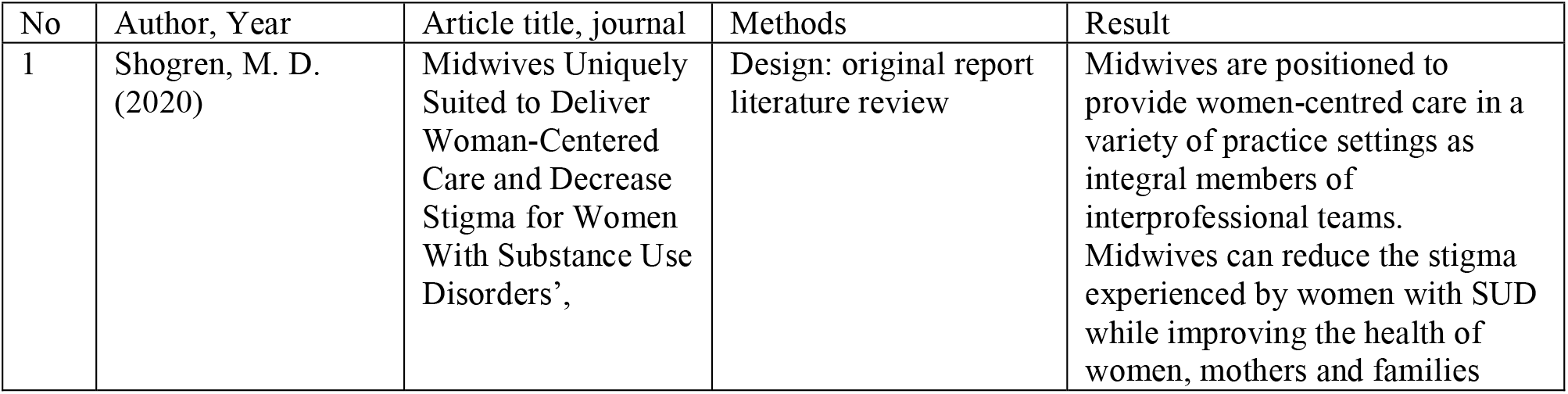
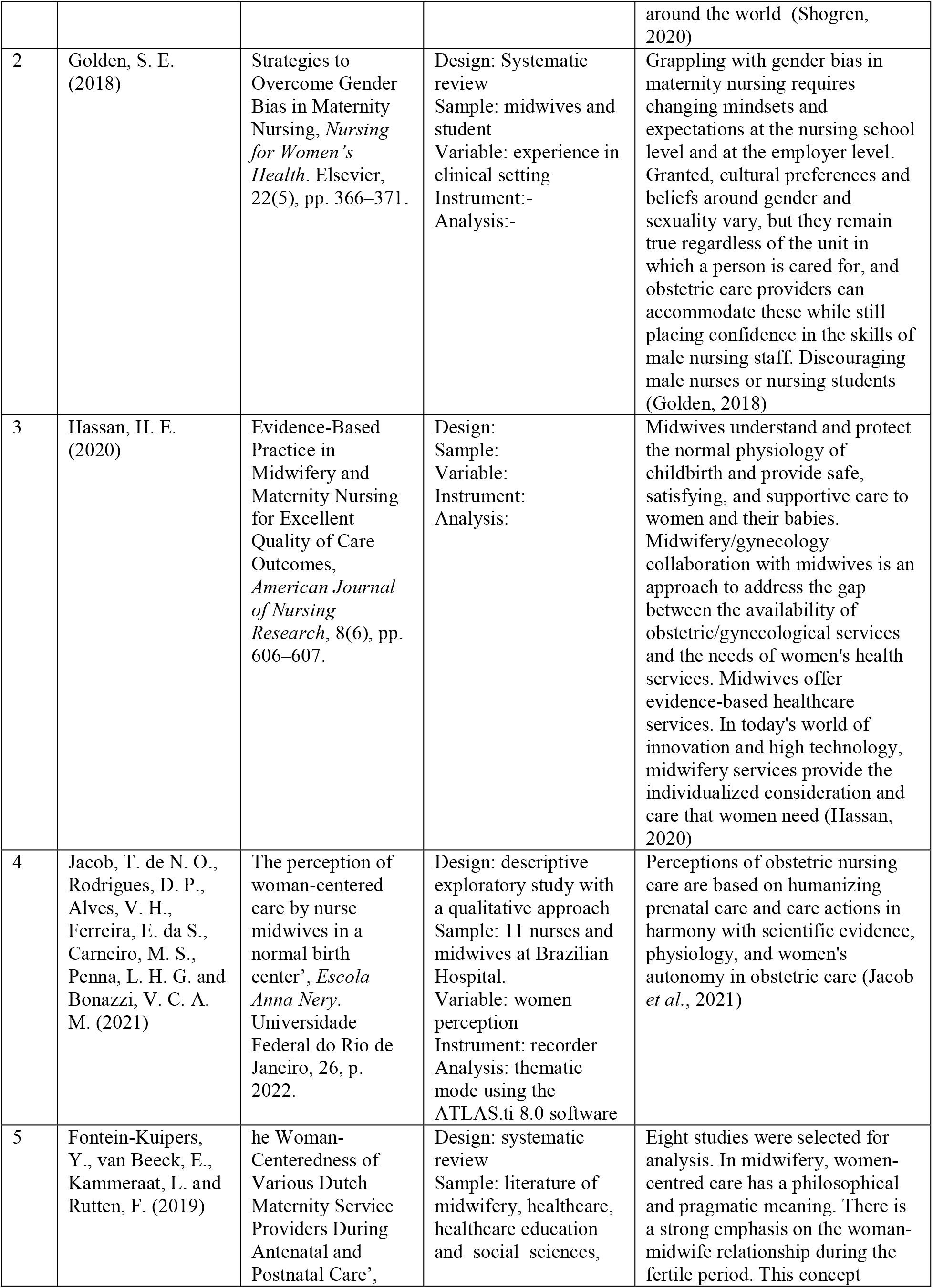
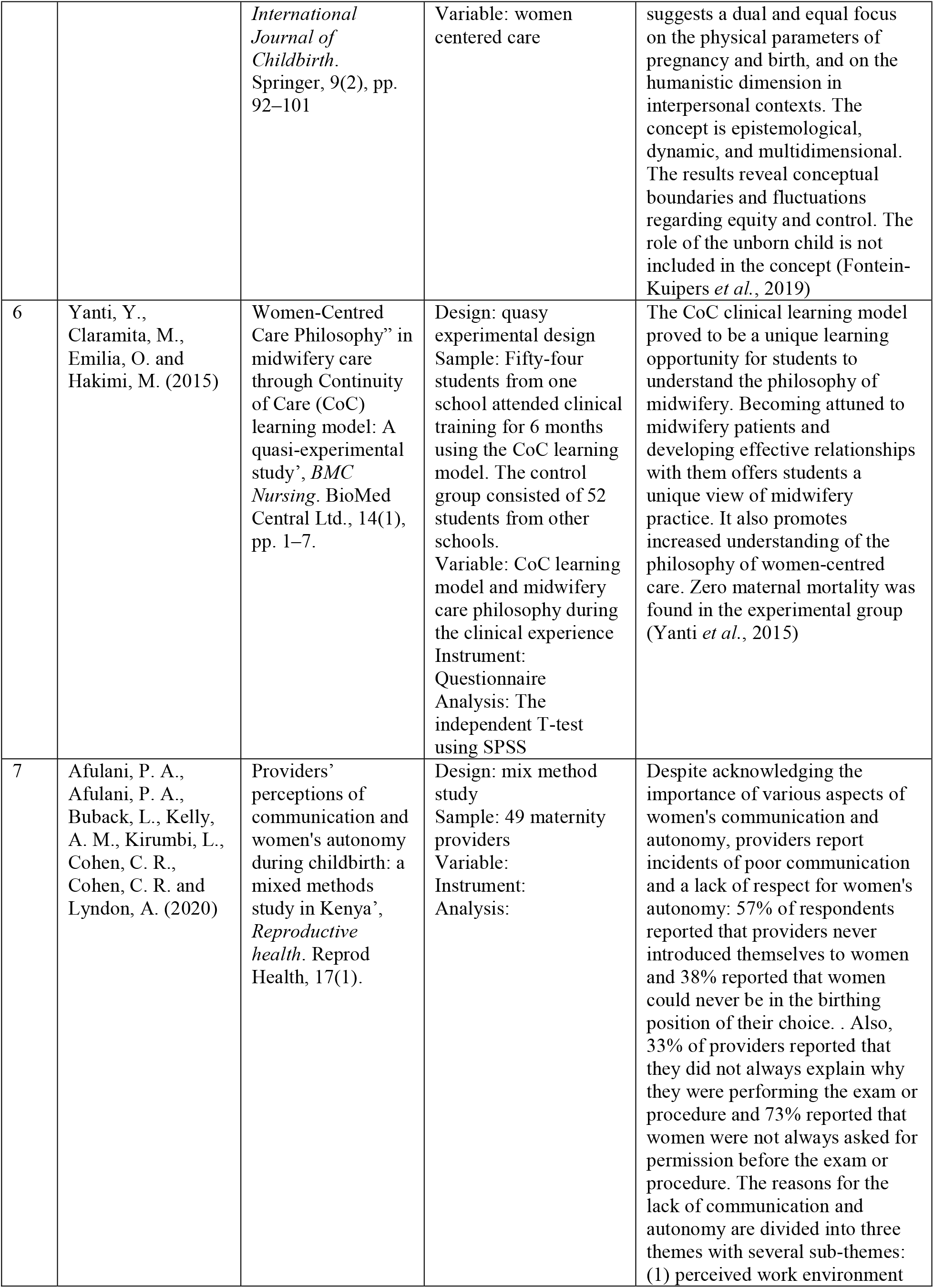
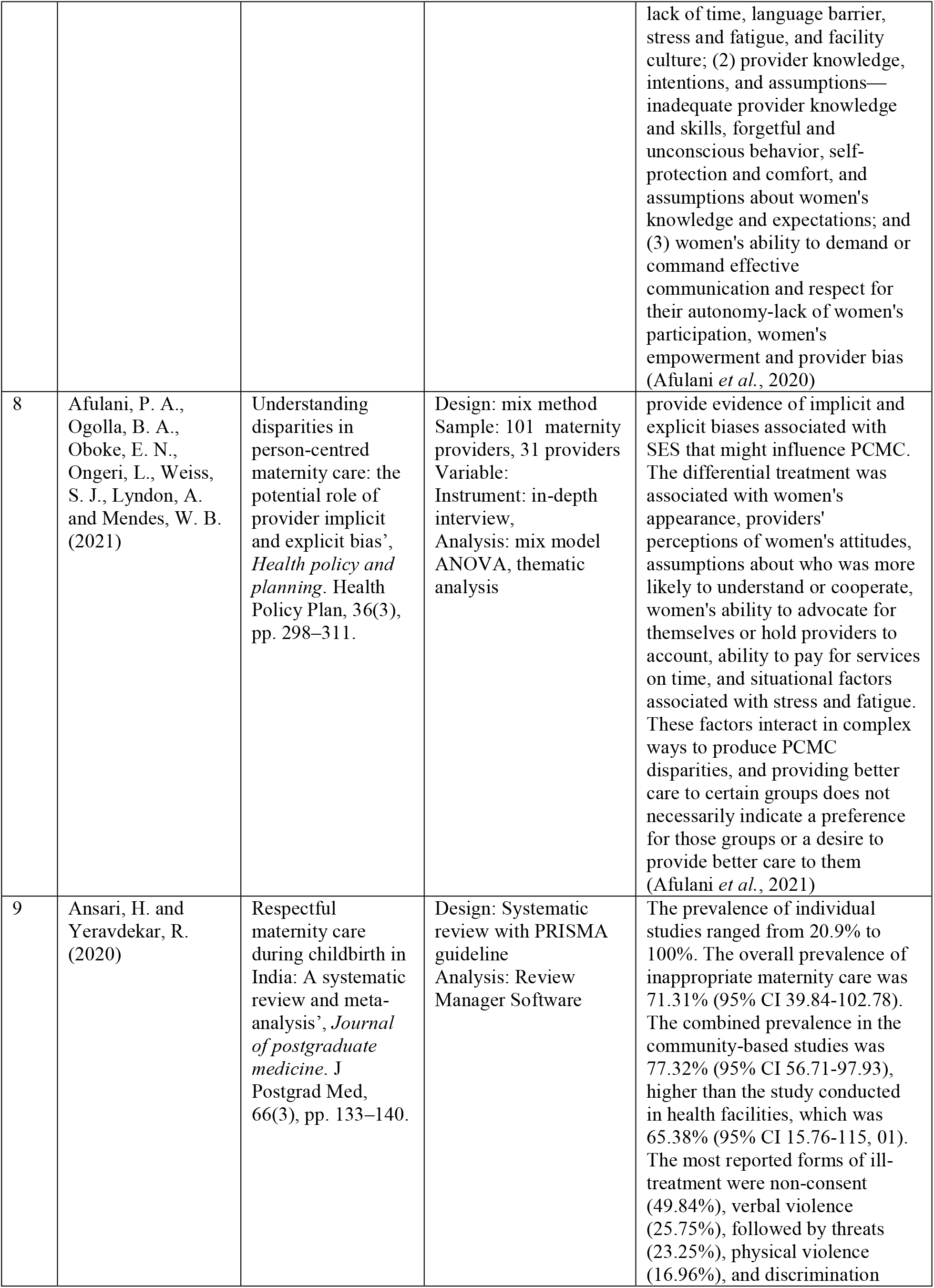
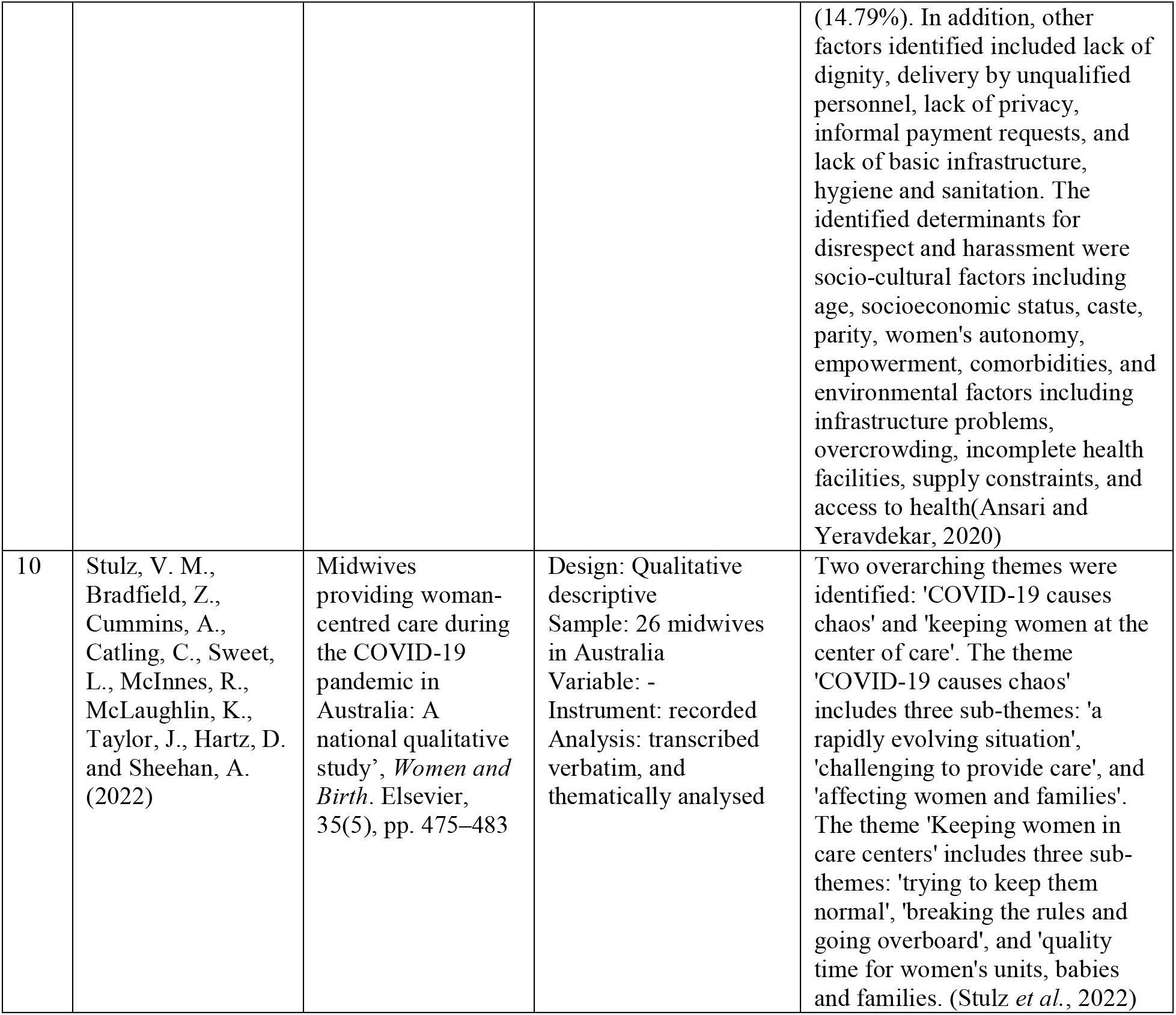
characteristics of reviewed articles

## 4. Discussion

Indonesia currently still has a high maternal mortality rate. Data The maternal mortality rate in 2017 was 177 per 100,000 live births while the SDG’s target by 2030 reduces the maternal mortality ratio to less than 70 per 100,000 live births (*SDG’s Dashboard*, no date). This requires the role of many parties in its completion, including the nursing profession. Maternity nursing as a maternal discipline, part of nursing is also required to provide nursing care in the prevention of mortality and morbidity in mothers and children.

Maternity nursing care is provided on an ongoing basis to women during pregnancy until delivery and newborns. Maternity nursing philosophy also believes that pregnancy and childbirth are not painful events, encourage increased spontaneous labor, reduce the occurrence of episiotomy and encourage exclusive breastfeeding. Maternity nursing care promotes, protects human beings, their reproductive, health and sexual rights, and respects ethnic and cultural diversity based on ethical principles of justice, harmony and respect for human dignity. Maternity nursing care is holistic and sustainable based on the scientific, social, emotional, cultural, spiritual, physical and psychological characteristics of women.

Maternity nursing care promotes care based on respect for human beings, compassion and human rights for every human being from the beginning of formation to the end of life. Maternity nursing care is also concerned with emancipation because it must protect and improve women’s health and status and build women’s confidence in their ability to give birth. Maternity nursing care takes place in partnership with women, recognizing their rights and respecting their determination in a continuous and non-authoritarian manner. Ethical and competent maternity nursing care must be accompanied by continuing education, scientific research and the application of empirical evidence.

The philosophy of maternity nursing views pregnancy and childbirth as normal and natural things so the participation and participation of families and communities are very much needed. Recent studies continue to develop when pregnant women do not only focus on the fetus or biomedical-clinical aspects but the mother on the psycho-social aspects. In addition, health education efforts in the community are also encouraged where women during pregnancy who become the center are a mother (Giarratano, 2003; Naughton, Harvey and Baldwin, 2021)so that the family, social environment, and health services (Bradfield *et al*., 2018)together provide support, pay attention to mothers both in terms of health, emotional needs and comfort when pregnant until giving birth. The results show that effective communication from health care providers and respect for women’s autonomy are important components of women-centered care (Compton *et al*., 2005). The results of research in Kenya are currently still lacking in women’s participation and women’s empowerment in making choices about health services (Afulani *et al*., 2020). Most women do not yet have the ability to demand or command effective communication and respect for their autonomy (Afulani *et al*., 2020).

Globally there are still many women who experience violence and neglect in health facilities when giving birth (Kassa, Tsegaye and Abeje, 2020). When women are pregnant and they also do not get the opportunity to make choices and services because they feel afraid, ashamed, and have low self-esteem toward health workers. It is a patient duty as a maternity nurse that every woman has the right to health services (Afulani *et al*., 2021).

Statistical data shows that an area in East Java, Indonesia has a fairly high maternal mortality rate even though the government has made various prevention efforts (Prasetyo *et al*., 2018). Maternity nursing science researches to find the cause with qualitative analysis of pregnant women, families, community leaders, and health workers. The results showed that when a pregnant woman gave birth with complications and an immediate referral to a higher health facility was required, the woman was not allowed to make decisions, but the extended family made the decision through a deliberation process. This turned out to be one of the causes of the high maternal mortality rate due to late referrals. The role of maternity nurses as advocates and educators collaborates with other disciplines to overcome this problem based on the results of the research.

In terms of family planning and the use of contraceptives in the community, most women cannot make a decision, but the decision rests with the husband. Whereas what we know, most contraceptives can used by women such as IUDs, injections, implants, and birth control pills. Women do not have freedom in terms of the right to their bodies and comfort. Many cases occur in the field when a husband does not allow his wife to use contraception even though each pregnancy is at high risk and causes the death of the mother and fetus. This is one of the reasons why maternal and infant mortality rates are still high.

In the health of the reproductive organs every woman is also at risk of experiencing disorders and diseases such as infections, sexually transmitted diseases, tumors, and malignancies. When women suffer from these diseases, both when they are girls and when they are married, it does not only have an impact on the physical but also psychological, because the reproductive function is related to sexual function which has to do with partners and basic human needs. Women who suffer and get social support will be at risk of having a body image disorder to low self-esteem and loss. In Indonesia, the second leading contributor to death in women is cervical cancer after breast cancer (Depkes RI, 2018). Maternity nursing in this case provides comprehensive nursing care including palliative care to improve the quality of life of patients.

## 5. Conclusion

Maternity nursing centers on women and pays attention to women and provides full service to women during pregnancy and childbirth and in childbearing age. Nurses and midwives as well as midwifery and nursing students need to know this to achieve a good quality of life for every woman

## Data Availability

All data produced in the present study are available upon reasonable request to the authors

## References

Afulani, P. A., Afulani, P. A., Buback, L., Kelly, A. M., Kirumbi, L., Cohen, C. R., Cohen, C. R. and Lyndon, A. (2020) ‘Providers’ perceptions of communication and women’s autonomy during childbirth: a mixed methods study in Kenya’, Reproductive health. Reprod Health, 17(1). doi: 10.1186/S12978-020-0909-0.

Afulani, P. A., Ogolla, B. A., Oboke, E. N., Ongeri, L., Weiss, S. J., Lyndon, A. and Mendes, W. B. (2021) ‘Understanding disparities in person-centred maternity care: the potential role of provider implicit and explicit bias’, Health policy and planning. Health Policy Plan, 36(3), pp. 298–311. doi: 10.1093/HEAPOL/CZAA190.

Ansari, H. and Yeravdekar, R. (2020) ‘Respectful maternity care during childbirth in India: A systematic review and meta-analysis’, Journal of postgraduate medicine. J Postgrad Med, 66(3), pp. 133–140. doi: 10.4103/JPGM.JPGM_648_19.

Bradfield, Z., Duggan, R., Hauck, Y. and Kelly, M. (2018) ‘Midwives being “with woman”: An integrative review’, Women and Birth. Elsevier, 31(2), pp. 143–152. doi: 10.1016/J.WOMBI.2017.07.011.

Compton, W. D., Fanjiang, G., Grossman, J. H., Reid, P. P., Institute of Medicine (U.S.), National Academies Press (U.S.) and National Academy of Engineering. (2005) Building a better delivery system a new engineering/health care partnership. National Academies Press. Available at: https://books.google.com/books/about/Building_a_Better_Delivery_System.html?hl=id&id=yvY34p56Ob4C (Accessed: 20 September 2022).

Depkes RI (2018) Riskesdas 2018.

Fontein-Kuipers, Y., van Beeck, E., Kammeraat, L. and Rutten, F. (2019) ‘The Woman-Centeredness of Various Dutch Maternity Service Providers During Antenatal and Postnatal Care’, International Journal of Childbirth. Springer, 9(2), pp. 92–101. doi: 10.1891/2156-5287.9.2.92.

Giarratano, G. (2003) ‘Woman-Centered Maternity Nursing Education and Practice’, The Journal of Perinatal Education. Lamaze International, 12(1), p. 18. doi: 10.1624/105812403X106694.

Golden, S. E. (2018) ‘Strategies to Overcome Gender Bias in Maternity Nursing’, Nursing for Women’s Health. Elsevier, 22(5), pp. 366–371. doi: 10.1016/J.NWH.2018.07.001.

Hassan, H. E. (2020) ‘Evidence-Based Practice in Midwifery and Maternity Nursing for Excellent Quality of Care Outcomes’, American Journal of Nursing Research, 8(6), pp. 606– 607. doi: 10.12691/ajnr-8-6-3.

Jacob, T. de N. O., Rodrigues, D. P., Alves, V. H., Ferreira, E. da S., Carneiro, M. S., Penna, L. H. G. and Bonazzi, V. C. A. M. (2021) ‘The perception of woman-centered care by nurse midwives in a normal birth center’, Escola Anna Nery. Universidade Federal do Rio de Janeiro, 26, p. 2022. doi: 10.1590/2177-9465-EAN-2021-0105.

Kassa, Z. Y., Tsegaye, B. and Abeje, A. (2020) ‘Disrespect and abuse of women during the process of childbirth at health facilities in sub-Saharan Africa: A systematic review and meta-analysis’, BMC International Health and Human Rights. BioMed Central Ltd, 20(1), pp. 1–9. doi: 10.1186/S12914-020-00242-Y/FIGURES/2.

Koblinsky, M., Moyer, C. A., Calvert, C., Campbell, J., Campbell, O. M. R., Feigl, A. B., Graham, W. J., Hatt, L., Hodgins, S. and Matthews, Z. (2016) ‘Quality maternity care for every woman, everywhere: a call to action’, The Lancet, 388(10057), pp. 2307–2320. Available at: http://ac.els-cdn.com/S0140673616313332/1-s2.0-S0140673616313332-main.pdf?_tid=bdc2b326-b936-11e6-bd0e-00000aab0f02&acdnat=1480755724_66dc6cbe97444e0aac893d33f9cdc47e.

Miller, S., Abalos, E., Chamillard, M., Ciapponi, A., Colaci, D., Comandé, D., Diaz, V., Geller, S., Hanson, C., Langer, A., Manuelli, V., Millar, K., Morhason-Bello, I., Castro, C. P., Pileggi, V. N., Robinson, N., Skaer, M., Souza, J. P., Vogel, J. P. and Althabe, F. (2016) ‘Beyond too little, too late and too much, too soon: a pathway towards evidence-based, respectful maternity care worldwide’, The Lancet. Elsevier, 388(10056), pp. 2176–2192. doi: 10.1016/S0140-6736(16)31472-6.

Naughton, S. L., Harvey, C. and Baldwin, A. (2021) ‘Providing woman-centred care in complex pregnancy situations’, Midwifery. Churchill Livingstone, 102, p. 103060. doi: 10.1016/J.MIDW.2021.103060.

Prasetyo, B., Damayanti, H. E., Pranadyan, R., Habibie, P. H., Romdoni, A. and Islami, D. (2018) ‘Maternal mortality audit based on district maternal health performance in East Java Province, Indonesia’, Bali Medical Journal (Bali Med J), 7(1), pp. 61–67. doi: 10.15562/bmj.v7i1.761.

SDG’s Dashboard (no date). Available at: http://sdgs-kesehatan.kemkes.go.id/index.php/sdgs/goal/3 (Accessed: 19 September 2022).

Shogren, M. D. (2020) ‘Midwives Uniquely Suited to Deliver Woman-Centered Care and Decrease Stigma for Women With Substance Use Disorders’, https://doi.org/10.1177/0844562120931663. SAGE PublicationsSage CA: Los Angeles, CA, 52(3), pp. 194–198. doi: 10.1177/0844562120931663.

Stockman, J. K., Syvertsen, J. L., Robertson, A. M., Ludwig-Barron, N. T., Bergmann, J. N. and Palinkas, L. A. (2014) ‘Women’s perspectives on female-initiated barrier methods for the prevention of HIV in the context of methamphetamine use and partner violence’, Women’s Health Issues. Division of Global Public Health, Department of Medicine, School of Medicine, University of California, La Jolla, San Diego, CA, United States: Elsevier USA, 24(4), pp. e397– e405. doi: 10.1016/j.whi.2014.04.001.

Stulz, V. M., Bradfield, Z., Cummins, A., Catling, C., Sweet, L., McInnes, R., McLaughlin, K., Taylor, J., Hartz, D. and Sheehan, A. (2022) ‘Midwives providing woman-centred care during the COVID-19 pandemic in Australia: A national qualitative study’, Women and Birth. Elsevier, 35(5), pp. 475–483. doi: 10.1016/J.WOMBI.2021.10.006.

Tunçalp, Were W. M., Maclennan, C., Oladapo, O. T., Gülmezoglu, A. M., Bahl, R., Daelmans, B., Mathai, M., Say, L., Kristensen, F., Temmerman, M. and Bustreo, F. (2015) ‘Quality of care for pregnant women and newborns—the WHO vision’, Bjog. Wiley-Blackwell, 122(8), p. 1045. doi: 10.1111/1471-0528.13451.

WHO recommendations on antenatal care for a positive pregnancy experience (no date). Available at: https://www.who.int/publications/i/item/9789241549912 (Accessed: 1 January 2021).

Yanti, Y., Claramita, M., Emilia, O. and Hakimi, M. (2015) ‘Students’ understanding of “Women-Centred Care Philosophy” in midwifery care through Continuity of Care (CoC) learning model: A quasi-experimental study’, BMC Nursing. BioMed Central Ltd., 14(1), pp. 1– 7. doi: 10.1186/S12912-015-0072-Z/TABLES/2.

